# Three-dimensional printing of patient-specific lung phantoms for CT imaging: emulating lung tissue with accurate attenuation profiles and textures

**DOI:** 10.1101/2021.07.30.21261292

**Authors:** Kai Mei, Michael Geagan, Leonid Roshkovan, Harold I. Litt, Grace J. Gang, Nadav Shapira, J. Webster Stayman, Peter B. Noël

## Abstract

**Purpose:** Phantoms are a basic tool for assessing and verifying performance in CT research and clinical practice. Patient-based realistic lung phantoms accurately representing textures and densities are essential in developing and evaluating novel CT hardware and software. This study introduces PixelPrint, a 3D printing solution to create patient-based lung phantoms with accurate attenuation profiles and textures.

**Methods:** PixelPrint, a software tool, was developed to convert Patient DICOM images directly into printer instructions (G-code). The density was modeled as the ratio of filament to voxel volume to emulate attenuation profiles for each voxel. A calibration phantom was designed to determine the mapping between filament line width and Hounsfield Units (HU) within the range of human lungs. For evaluation of PixelPrint, a phantom based on a human lung slice was manufactured and scanned with the same CT scanner and protocol used for the patient scan. Density and geometrical accuracy between phantom and patient CT data was evaluated for various anatomical features in the lung.

**Results:** For the calibration phantom, measured mean Hounsfield units show a very high level of linear correlation with respect to the utilized filament line widths, (r > 0.999). Qualitatively, the CT image of the patient-based phantom closely resembles the original CT image both in texture and contrast levels, with clearly visible vascular and parenchymal structures. Regions-of-interest (ROIs) comparing attenuation illustrated differences below 15 HU. Manual size measurements performed by an experienced thoracic radiologist reveal a high degree of geometrical correlation of details between identical patient and phantom features, with differences smaller than the intrinsic spatial resolution of the scans.

**Conclusion:** The present study demonstrates the feasibility of 3D printed patient-based lung phantoms with accurate organ geometry, image texture, and attenuation profiles. PixelPrint will enable applications in the research and development of CT technology, including further development in radiomics.

## Introduction

Anthropomorphic phantoms, geometric image quality phantoms, and mathematical phantoms are fundamental tools for developing, optimizing, and evaluating novel methods in computed tomography (CT) research and clinical practice. Common CT phantoms are typically manufactured by casting, forming, and molding homogenous materials such as resin or plastic. While many different phantoms are available commercially and in research laboratories, there is a lack of patient-based phantoms that fully represent attenuation profiles and textures seen in clinical CT acquisitions, for example, for healthy and diseased lungs. Additionally, the academic and clinical CT community would benefit from a rapid and inexpensive manufacturing process compared to current commercial solutions.

Over the last decade, fused deposition modeling (FDM)-based 3-dimensional (3D) printing of various tissue-mimicking phantoms has been widely explored for validation and evaluation of CT imaging technology^1–6^. Studies have focused on several areas, including manufacturing geometrically correct models of organs^7–11^, generating realistic texture samples^12–14^, and creating accurate attenuation profiles^15–18^. The general procedure to 3D print an anthropomorphic phantom from CT image data includes: (i) segmentation of regions/organs of interest in CT images, (ii) conversion of selected regions from volumetric data to triangulated surface geometry models (e.g., STL or SLA files), and (iii) use of printer-specific slicing software to apply proper parameters (e.g., extrusion rate, print speed, infill ratios, etc.) and generate instructions (G-code) for printers to create 3D products. While this approach produces phantoms that better resemble true anatomical structures, it still has shortcomings. First, spatial resolution is largely lost due to segmentation of regions and conversion to surface models. Second, for each region/surface model, the slicer software assigns unique infill and exterior walls (or perimeter), creating abrupt, unrealistic transitions between regions of different densities in the final product. Third, due to its reliance on segmentation, this method is susceptible to boundary placement errors.

A promising alternative is to directly translate DICOM image data into G-code^17,18^. To generate different densities in 3D printed CT phantoms, these methods utilize a pixel-by-pixel change in the filament extrusion rate, while maintaining a constant printing speed. Although this approach enables generation of sophisticated phantoms with realistic attenuation profiles, it falls short when printing high-resolution features. This reduction in spatial resolution is an important concern when generating natural image textures. Therefore, there is an unmet need for a quick and cost-efficient process for generating patient-based phantoms with accurate organ geometry, image texture, and attenuation profiles.

We propose a 3D printing solution that is capable of achieving accurate organ geometry, image texture, and attenuation profiles while eliminating the complexities and limitations of previous methods. Our solution is a one-step method for translating CT images into printer instructions (G-code) that can be used by any FDM 3D printer. It combines varying printer speeds with a constant filament extrusion rate to control the density of each printed voxel. In the following sections we present a complete description of the proposed method, as well as results from successful proof-of-principle experiments with geometrical and patient-based lung phantoms.

## Materials and Methods

### PixelPrint

Conventional 3D printing utilizes slicing software to convert 3D models (e.g., STL files) to printer instructions written in G-code, a widely used machine language defining 3D printing parameters (e.g., layer height, retraction, print speed, etc.). We present a solution that accepts volumetric CT DICOM data as input and converts these data directly into G-code without segmentation or intermediate 3D models.

Applied to common FDM 3D printers, PixelPrint produces multiple 2D layers, one layer at a time, to create 3D objects or phantoms. Each printed layer is mapped from the corresponding DICOM slice, with the physical scale controlled to ensure that the resulting phantom has the same dimensions as the scanned patient. For each layer in the 3D printed phantom, PixelPrint generates an array of spaced, parallel filament lines. The lines are at fixed spacing but are of varying widths, creating a partial volume effect to form varying densities in the CT scan, i.e., wider line widths for high-density areas and narrower line widths for low-density areas. PixelPrint computes the density of the input image at closely spaced intervals along each line and maps it into appropriate extrusion and printhead speeds over each interval. It then records one G-code command that defines a starting point, an end point, the filament extrusion, and speed for that interval. This process is repeated for every interval over every line, in every layer, until the whole phantom is fully encoded in a G-code file. Since layers are deposited in alternating directions, the varying line widths create a matrix of high- and low-density regions that correlate with the original 3D input image volume. The matrices are shifted in angle and location at each layer, and the printed layer height is much smaller than the typical CT slice thickness. Each CT slice of the printed phantom will therefore contain multiple shifted layers, ensuring that reslicing of the CT image data will not result in sampling or moiré patterns.

In our experiments, we found that altering the line width by varying the extrusion rate alone does not provide sufficient spatial resolution due to the inherently slow response time of the extrusion process. Instead, we maintain a constant filament flow rate while changing the speed of the printhead to control the extrusion width.

### Phantom Design

#### Calibration Phantom

The partial volume effect created by varying the filament line width with fixed line spacing determines the (local) mean material density and thus the x-ray attenuation in a CT scan. A calibration phantom was designed to determine the mapping between filament line widths and Hounsfield Units (HU) within the range of human lungs. A multi-sector phantom (cylinder divided into radial slices) with a diameter of 100 mm and a height of 10 mm was printed using PixelPrint. The phantom comprises ten sections with different material densities, from 10% to 100% at 10% intervals (Figure 1). CT measurements of this phantom were used to determine the 3D printed maximum and minimum HU of PixelPrint, as well as the conversion between HU and filament line widths for patient-based phantoms.

**Figure 1.**
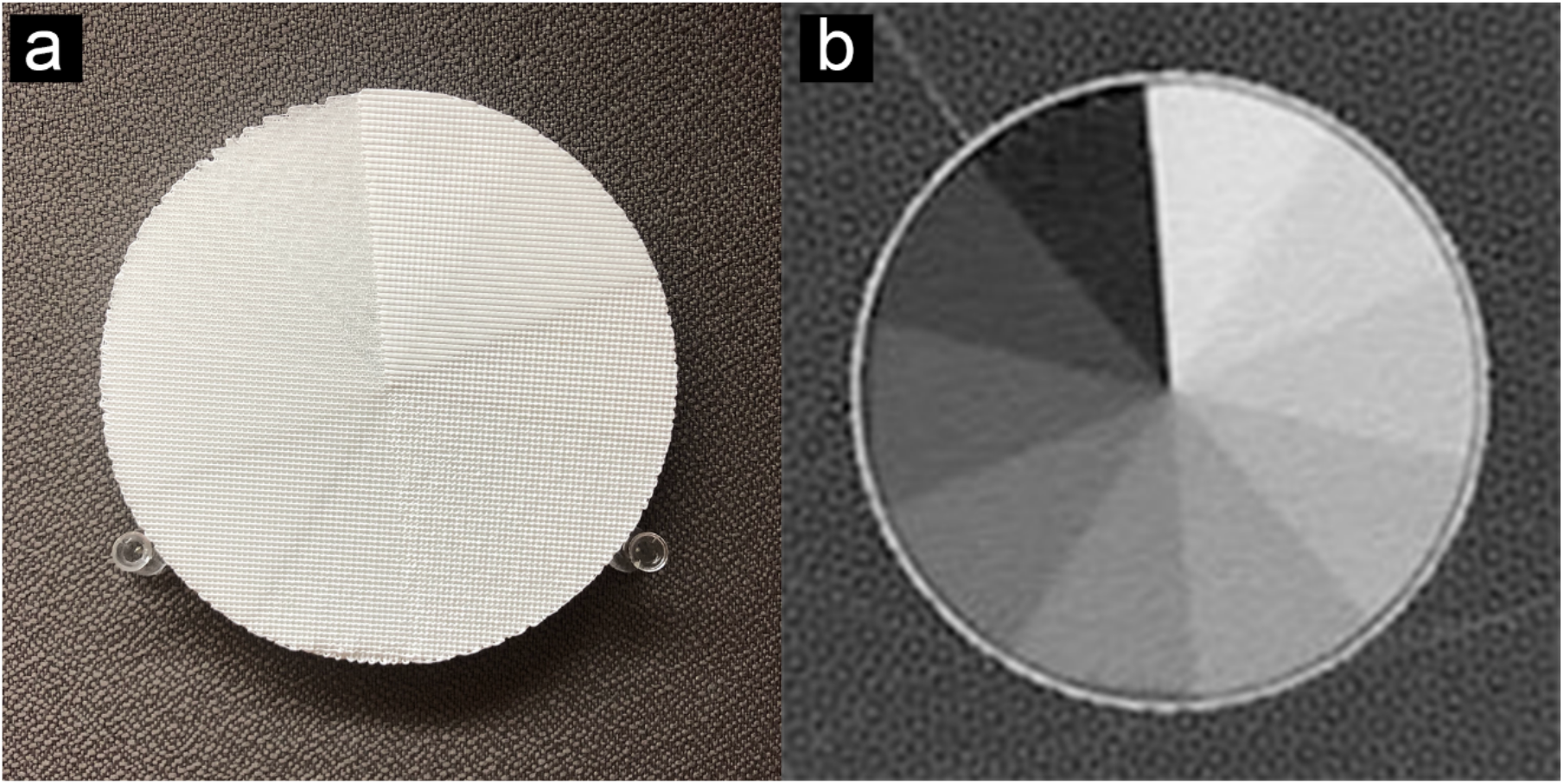
Calibration phantom. (a) Photo of the multi-sector calibration phantom. (b) CT image of the phantom. Window level is -400 HU. Window width is 1500 HU.

#### Patient-based Phantom

The Institutional Review Board (IRB) approved this retrospective study. A single data set of a patient who had been diagnosed with COVID-19 pneumonia and acute respiratory distress syndrome (ARDS) was selected from the PACS system at the Hospital of the University of Pennsylvania and anonymized. The CT images demonstrate extensive fibro-proliferative changes with both interstitial and alveolar components throughout the lung parenchyma. Imaging was performed in the supine position on a dual-source CT scanner (Somatom Drive, Siemens Healthineers, Erlangen, Germany). Table 1 presents acquisition and reconstruction parameters utilized for imaging. Since this proof-of-principle study focuses on lung imaging, the right lung of the patient was selected as input to reduce printing time and complexity. A 20 mm diameter ring surrounding the lung was added for better positioning of the phantom within the bore of a 300 x 400 mm^2^ oval phantom representing a medium-sized patient (see details below). HU values were converted into filament line widths using the mapping calculated from the calibration phantom described above. A lower cut-off value of 10% and an upper cap of 100% material density were applied. The phantom is shown in Figure 3.

**Table 1.** Scan protocol for the patient-based phantom

|  |  |
| --- | --- |
| Scanner model | Siemens SOMATON Drive |
| Tube voltage | 100 kVp (single source mode) |
| Tube current (at slice position) | 292 mA |
| Rotation time | 0.5 seconds |
| Spiral pitch factor | 1.2 |
| Exposure (at slice position) | 121 mAs |
| CTDI<sub>vol</sub> (at slice position) | 4.54 mGy |
| Slice thickness | 1 mm |
| Reconstruction filter | Br49f(3) (diagnostic lung sharpest with iterative reconstruction ADMIRE level 3) |
| Reconstructed field of view | 404 x 404 mm<sup>2</sup> |
| Matrix size | 512 x 512 pixel<sup>2</sup> |
| Pixel spacing (in x and y) | 0.79 mm |

### Phantom Production

Phantoms were printed on a fused-filament 3D printer (Lulzbot TAZ 6 with M175 tool head, Fargo Additive Manufacturing Equipment 3D, LLC Fargo, ND, USA) using a 0.25 mm brass nozzle. Polylactic Acid (PLA) filament with a diameter of 1.75 mm (MakeShaper, Keene Village Plastics, Cleveland, OH, USA) was extruded at a nozzle temperature of 210 °C. To improve adhesion, the build plate was heated to 50 °C. Printing speed varied from 3.0 to 30 mm/s, producing line widths from 1.0 to 0.1 mm.

### Data Acquisition & Analysis

For imaging, printed phantoms were placed inside the 20 cm bore of a technical phantom (Gammex multi-energy CT phantom, Sun Nuclear Corporation, Melbourne, FL, USA) to mimic attenuation profiles of an average-sized patient (300 x 400 mm^2^). Imaging was performed with the same CT scanner using the same protocol as the clinical acquisition (Table 1).

The scan of the multi-sector calibration phantom was performed with a higher tube current (800 mA) to reduce the effects of Poisson noise. Square regions-of-interest (ROI) with a fixed size of 14 x 14 pixels were manually positioned on the CT image to calculate HU statistics within the ten sectors of 10% to 100% material densities. Mean HU values and standard deviations were measured and Pearson’s correlation coefficient was calculated using linear regression.

The scan of the patient-based phantom was performed with the same radiation dose level used for the patient scan (CTDIvol of 4.54 mGy at slice position). The resulting image was exported and registered to the original patient image using an affine transform available with the OpenCV library^19^. ROIs of different sizes in varied locations were manually placed in the vessel and parenchymal areas by an experienced thoracic radiologist (L.R., four years of experience) using ImageJ (National Institutes of Health, USA). See Figure 4 for exact ROI positions. Mean HU values and standard deviations were compared between the patient DICOM image and the phantom DICOM image. In addition, manual size measurements of three small oval structures were performed by the radiologist on both patient and phantom images using RadiAnt DICOM viewer (Medixant, Poznań, Poland).

## Results

Printing of the multi-sector calibration phantom required 16 hours of printing time. The CT image of the phantom shows homogeneous appearances within the ten different material density regions. Figure 1 illustrates that individual printed filament lines are not visible. Measured mean HU values show a very high level of linear correlation with respect to the utilized filament line widths, with a Pearson’s correlation coefficient r > 0.999 (see Figure 2). The maximum HU (corresponding to 100% material density) was measured as 115 and the minimum HU (corresponding to 10% material density) was measured as -867.

**Figure 2.**
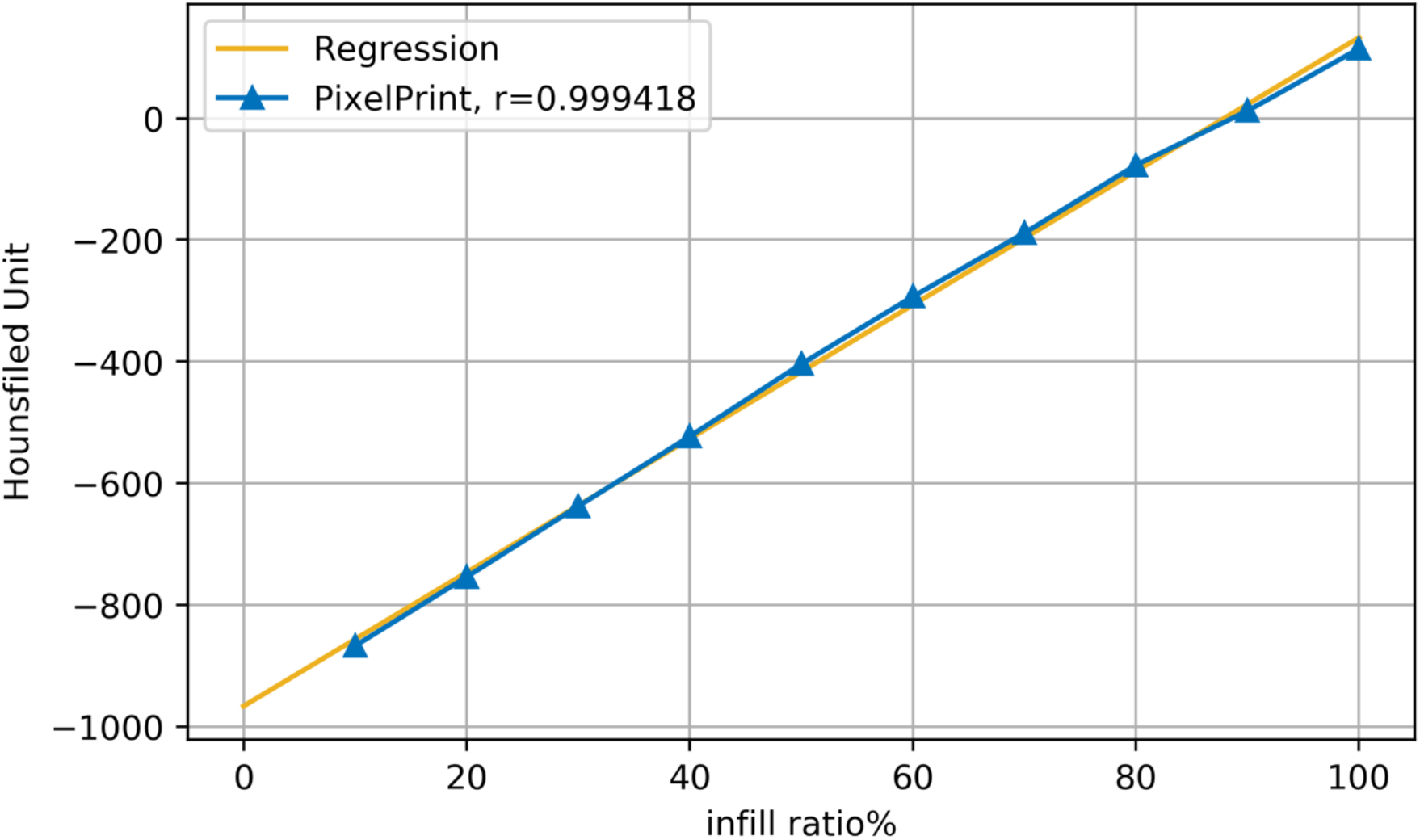
Linear regression between material density and Hounsfield Units. Blue triangles show the mean values for regions of material density 10% to 100% in the multi-sector calibration phantom. The yellow line represents the regression line. r is Pearson’s correlation coefficient.

The results from the patient-based phantom are summarized in Figure 3. Panels a, b, and c show a photograph of the phantom, CT slice of the phantom, and original patient data, respectively. Printing of the patient-based phantom required 24 hours of printing time. The CT image of the phantom, shown in zoomed-in regions in the lower panels in Figure 3, closely resembles the original CT image in both texture and contrast levels, with clearly visible vascular and bronchial structures. Figure 4 shows the identical regions in patient and phantom data selected for density measurements. Although the patient image appears noisier than the phantom image, due to higher attenuation from the patient body, five ROIs show very similar mean values, with differences less than 15 HU (see Table 2). In general, lower density areas have a slightly higher difference due to the 10% material density cut-off used with PixelPrint.

**Figure 3.**
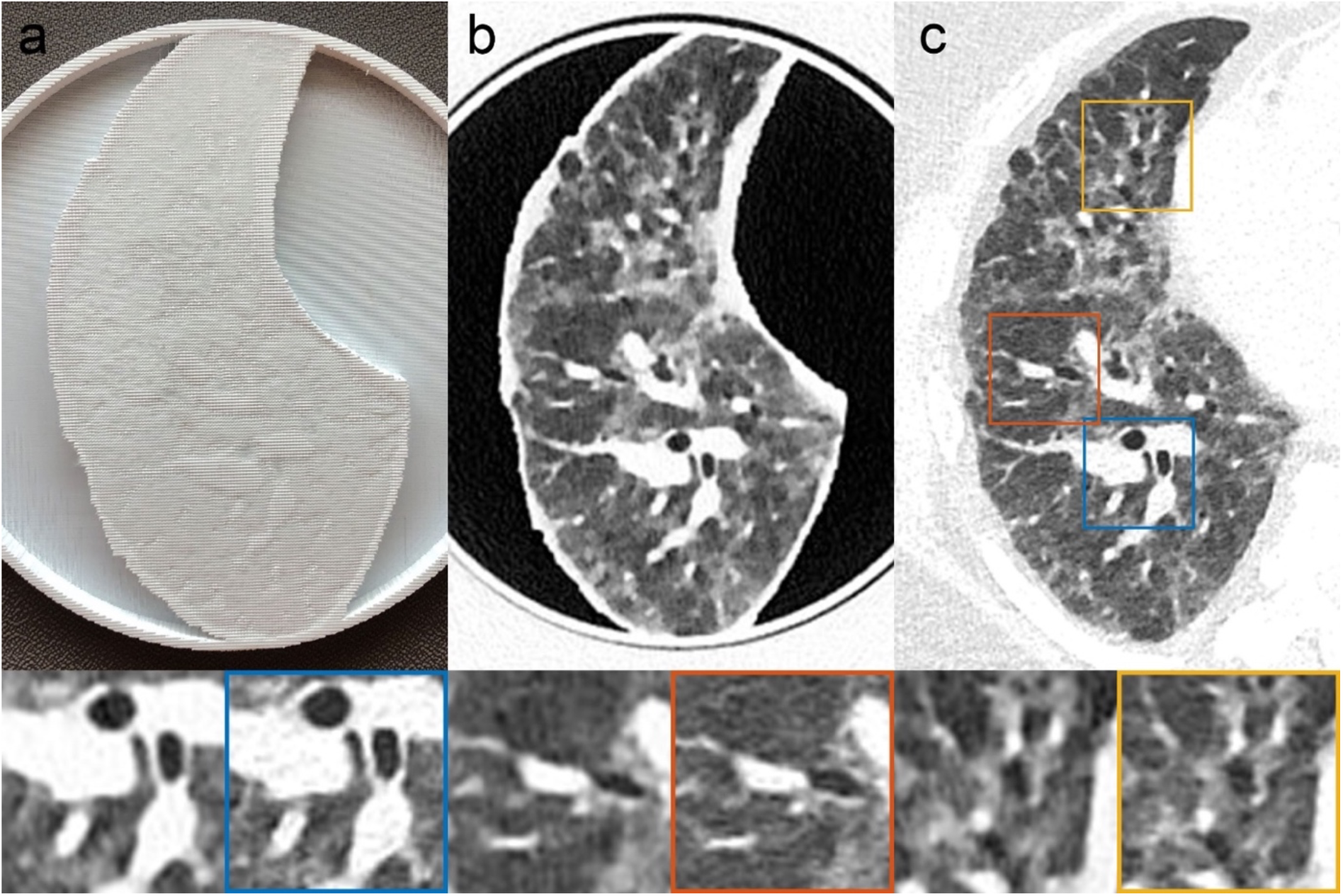
Patient-based Lung Phantom visually highly resembles the original CT image both in texture and contrast levels. (a) Photography of the printed patient-based phantom. (b) CT image of patient-based phantom. (c) CT image of patient lung. Yellow, red and blue boxes indicate zoomed-in regions of the patient DICOM image. Window level is -500 HU. Window width is 1000 HU.

**Figure 4.**
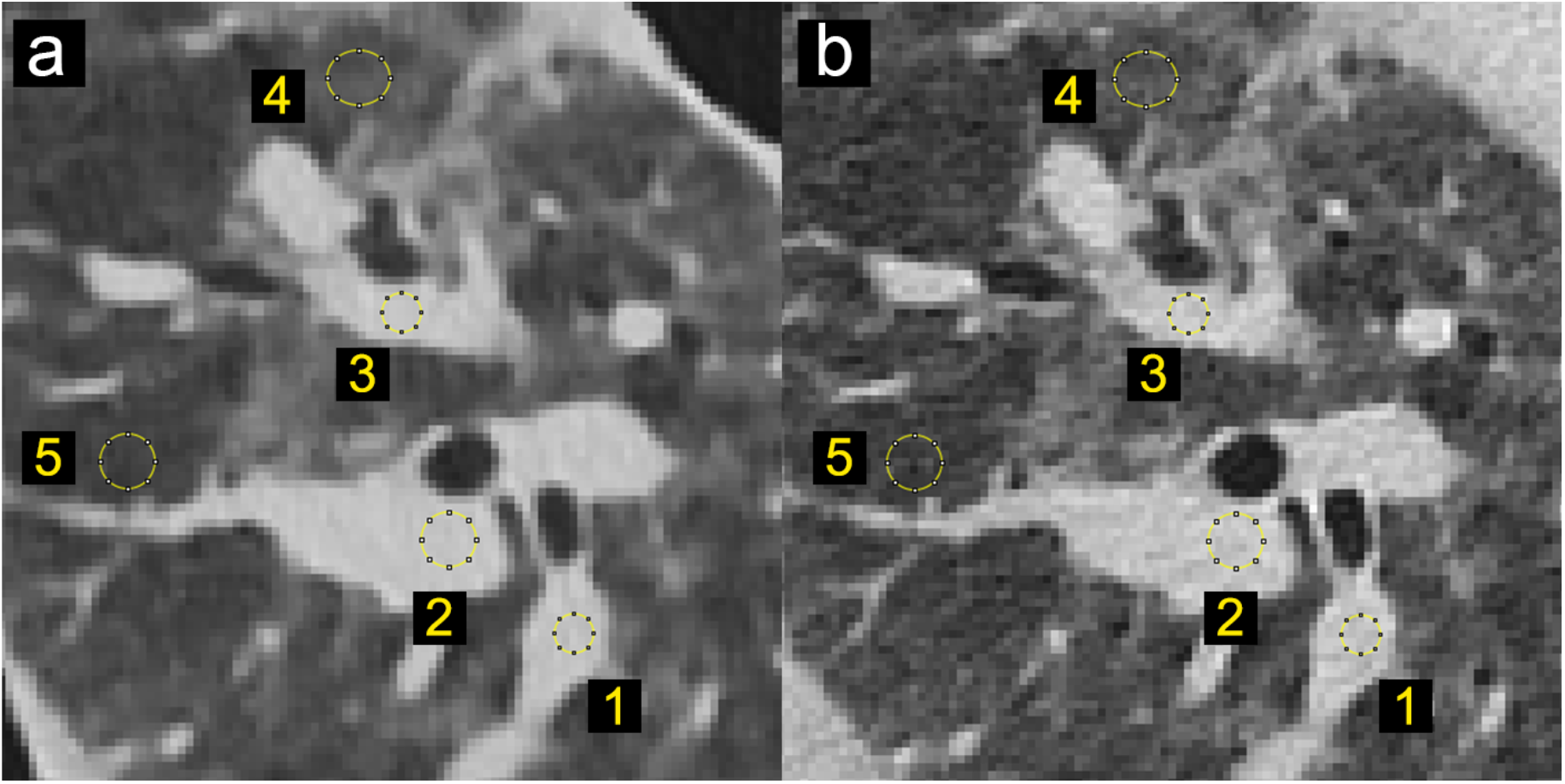
Locations and size of the selected regions of interest for density measurements in patient and phantom data. (a) CT image of patient-based phantom. (b) CT image of original patient lung. Window level is -500 HU. Window width is 1000 HU.

**Table 2.**
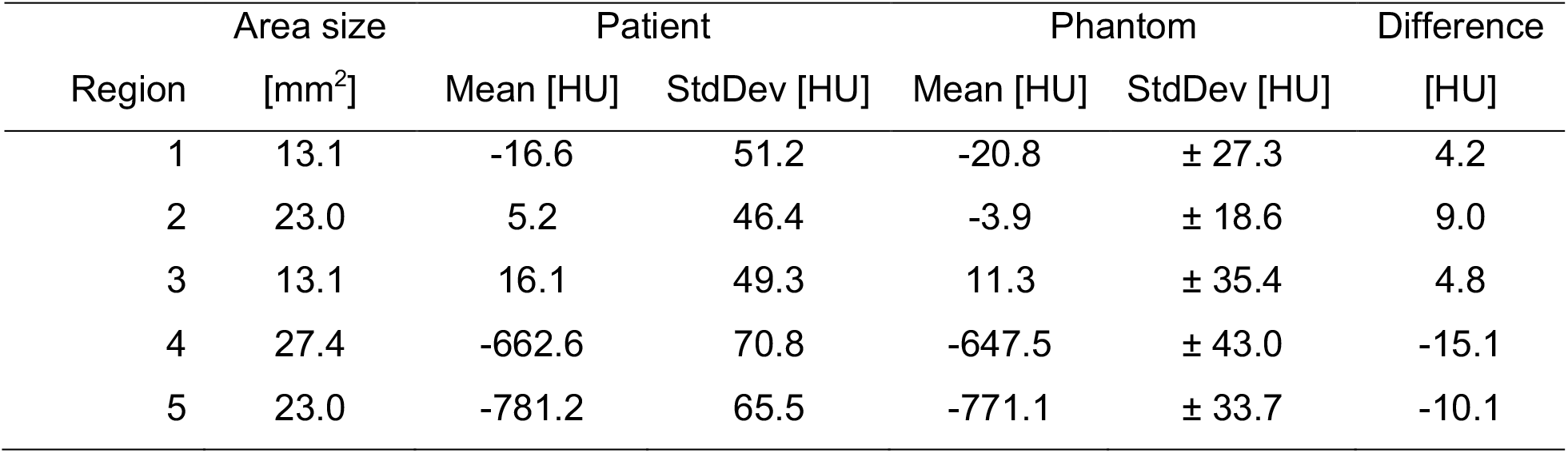
Mean and standard deviation of density measurements in patient and phantom data.

| Region | Area size<br>[mm<sup>2</sup>] | Patient |  | Phantom |  | Difference<br>[HU] |
| --- | --- | --- | --- | --- | --- | --- |
|  |  | Mean [HU] | StdDev [HU] | Mean [HU] | StdDev [HU] |  |
| 1 | 13.1 | -16.6 | 51.2 | -20.8 | ± 27.3 | 4.2 |
| 2 | 23.0 | 5.2 | 46.4 | -3.9 | ± 18.6 | 9.0 |
| 3 | 13.1 | 16.1 | 49.3 | 11.3 | ± 35.4 | 4.8 |
| 4 | 27.4 | -662.6 | 70.8 | -647.5 | ± 43.0 | -15.1 |
| 5 | 23.0 | -781.2 | 65.5 | -771.1 | ± 33.7 | -10.1 |

Figure 5 presents the three manually measured anatomical features. Manual size measurements performed by the radiologist illustrate a high degree of geometrical correlation of details between the patient image and the phantom images, with differences smaller than the intrinsic spatial resolution of the scans (see Table 3).

**Figure 5.**
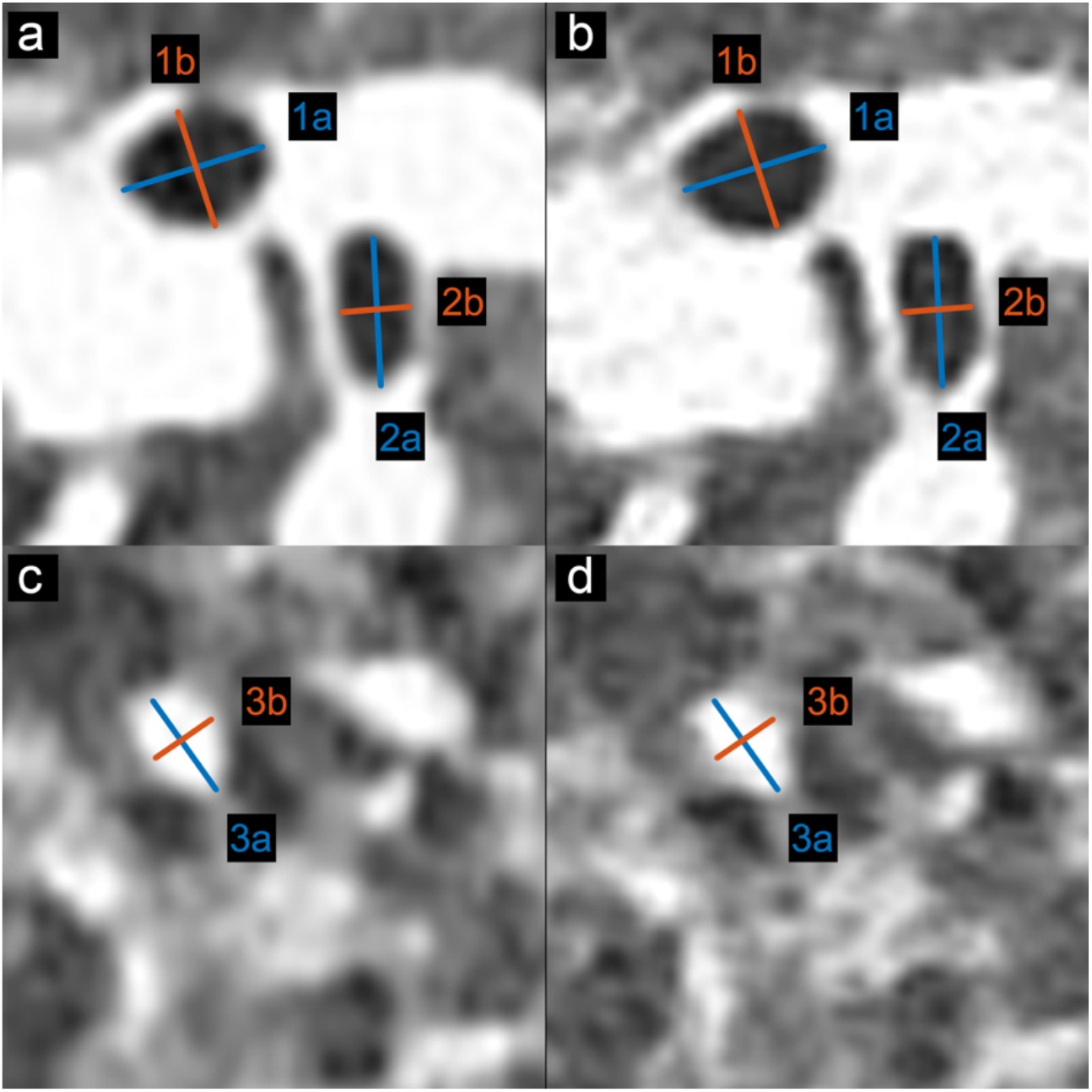
Locations and size of the selected anatomical features for size measurements in patient and phantom data. (a) and (c) CT image of patient-based phantom. (b) and (d) CT image of original patient lung. Window level is -500 HU. Window width is 1000 HU.

**Table 3.**
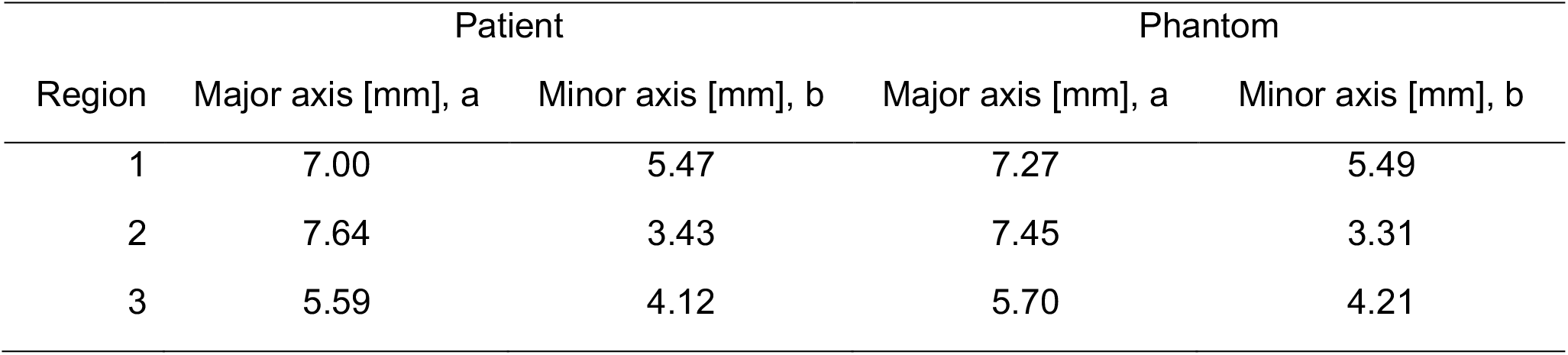
Size measurement of anatomical features in patient and phantom data.

| Region | Patient |  | Phantom |  |
| --- | --- | --- | --- | --- |
|  | Major axis [mm], a | Minor axis [mm], b | Major axis [mm], a | Minor axis [mm], b |
| 1 | 7.00 | 5.47 | 7.27 | 5.49 |
| 2 | 7.64 | 3.43 | 7.45 | 3.31 |
| 3 | 5.59 | 4.12 | 5.70 | 4.21 |

## Discussion

This paper introduces a novel 3D printing method, PixelPrint, that allows generation of patient-based phantoms with accurate organ geometry, image texture, and attenuation profiles. This method allows direct conversion of CT DICOM images into 3D printer instructions without segmentation. By eliminating segmentation, PixelPrint produces smooth, lifelike transitions between regions of different density. PixelPrint produces no boundary effects. The algorithm is strictly analytical with no filtering and no tuning parameters. The one-step translation of image data reduces information loss (due to segmentation and triangulation steps in the 3D modeling) and maintains spatial resolution. Regarding resolution, our variable speed printing concept allows us to control density on a voxel-by-voxel basis. Thus, we are enabling a cost-effective manufacturing process that generates realistic models of human lungs without sacrificing spatial and contrast resolution.

Over the last decade, several approaches have been proposed to produce clinically applicable CT phantoms. Kairn *et al*. introduced a method to generate a patient-based lung phantom^20^. They segmented CT images of the lung into three different regions and produced a tissue equivalent lung phantom. However, their approach is not able to meet the resolution requirements to represent structures in the lung parenchyma. Giron *et al*. and Joemai *et al*. developed a printed lung for image quality assessment in CT; their prints contain vascular structures with limited realistic lung textures^10,21^. Okkalidis *et al*. proposed a pixel-by-pixel algorithm^14,17^, translating DICOM images to printer instructions and printed patient-specific skull and chest phantoms. Results showed a reliable match in HU; however, detailed structures and textures within the lung are not visible. Jahnke *et al*. also introduced an alternative approach^22,23^ of stacking radiopaque 2D prints to form patient-based 3D phantom.

Our method enables the creation of real ground truth from clinical CT data, opening opportunities in the clinical and research arena. For day-to-day operations, our phantom concepts allow optimizing CT protocols with a focus on specific clinical tasks. For example, the clinical introduction of advanced non-linear reconstruction algorithms^24^ can be challenging due to the limited clinical value of technical phantoms and ethical difficulties of scanning patients twice for this purpose. With our phantoms, an ample parameter space can be evaluated to determine the optimal solution with respect to radiation exposure and diagnostic image quality. A positive effect could be achieved for CT research and development by accelerating clinical evaluations with patient-based phantoms. Predominantly novel data-driven developments in artificial intelligence and radiomics can gain significantly from early access to realistic clinical data. One open challenge is the effect of differences in CT protocols and inter-vendor variabilities on radiomic features^25–28^. With a representative group of patient-based phantoms manufactured with PixelPrint, one would be able to evaluate this effect fully and determine a robust and rigorous operating space for radiomic feature extraction. Further, the same group of phantoms may assist as a tool to evaluate and validate harmonization strategies.

The present study has some limitations. Only one patient-based phantom was evaluated for this proof-of-concept study. Follow-up studies will provide additional data and measurement to describe specific lung diseases such as COVID-19 pneumonia. For our initial study, we have focused on generated realistic models of the human lung. Future studies will be essential to add the capability to print soft tissue and bone to cover a broader range of anatomical regions and tissue x-ray attenuation.

Finally, we would like to allow the larger medical, academic, and industrial CT community to have access to PixelPrint. We can make copies of our phantoms available as well as customized phantoms based on specific CT images, which could include various lung diseases. For additional information, please see the project homepage: www.pennmedicine.org/CTResearch/PixelPrint.

## Conclusion

In conclusion, the present study is the first to illustrate the possibility of creating 3D printed patient-based lung phantoms with accurate organ geometry, image texture, and attenuation profiles. This may lead to a paradigm change for the development of novel CT hardware and software by enabling accelerated evaluation and validation with realistic patient-based data. Ultimately this will shape the clinical day-to-day routine and benefit patients with novel and standardized CT imaging.

## Acknowledgement

We acknowledge support through the National Institutes of Health (R01CA249538).

